# Untargeted saliva metabolomics reveals COVID-19 severity

**DOI:** 10.1101/2021.07.06.21260080

**Authors:** Cecile F. Frampas, Katie Longman, Matt P. Spick, Holly M. Lewis, Catia D. S. Costa, Alex Stewart, Deborah Dunn-Walters, Danni Greener, George E. Evetts, Debra Skene, Drupad Trivedi, Andrew R. Pitt, Katherine Hollywood, Perdita Barran, Melanie J. Bailey

## Abstract

**Background:** The COVID-19 pandemic is likely to represent an ongoing global health issue given the potential for vaccine escape and the low likelihood of eliminating all reservoirs of the disease. Whilst diagnostic testing has progressed at pace, there is an unmet clinical need to develop tests that are prognostic, to triage the high volumes of patients arriving in hospital settings. Recent research has shown that serum metabolomics has potential for prognosis of disease progression. ^1^ In a hospital setting, collection of saliva samples is more convenient for both staff and patients, and therefore offers an alternative sampling matrix to serum. We demonstrate here for the first time that saliva metabolomics can reveal COVID-19 severity.

**Methods:** 88 saliva samples were collected from hospitalised patients with clinical suspicion of COVID-19, alongside clinical metadata. COVID-19 diagnosis was confirmed using RT-PCR testing. COVID severity was classified using clinical descriptors first proposed by SR Knight et al. Metabolites were extracted from saliva samples and analysed using liquid chromatography mass spectrometry.

**Results:** In this work, positive percent agreement of 1.00 between a PLS-DA metabolomics model and the clinical diagnosis of COVID severity was achieved. The negative percent agreement with the clinical severity diagnosis was also 1.00, for overall percent agreement of 1.00.

**Conclusions:** This research demonstrates that liquid chromatography-mass spectrometry can identify salivary biomarkers capable of separating high severity COVID-19 patients from low severity COVID-19 patients in a small cohort study.

## 1. Introduction

The SARS-CoV-2 pandemic has caused a sustained threat to global health since the discovery of the virus in 2019. ^2^ Whilst great strides have been made in both treatment and vaccination development, ^3,4^ the disease has inflicted multiple waves of infection throughout the world during 2020 and into 2021. ^5,6^ COVID-19 has higher fatality rates than seasonal influenza, ^7^ and in addition, new variants are constantly evolving with the potential for either reduced vaccine effectiveness or altered lethality. ^8^ As a consequence, there is a continuing need both for better understanding of the impact of COVID-19 on the host metabolism as well as for prognostic tests that can be used to triage the high volumes of patients arriving in hospital settings.

Nasopharyngeal swabs followed by polymerase chain reaction (PCR) have been adopted worldwide for SARS-CoV-2 detection. However, supply chains for swabs rapidly collapsed amongst exponential increases in demand for testing, highlighting the urgency for alternative sample types and testing approaches. Furthermore, whilst PCR tests are easily deployable and highly selective for the virus, these approaches yield no prognostic information and cannot easily be delivered for rapid turnaround at the point of care, for example during a hospital admissions process. In contrast, tests based on mass spectrometry can be provided in minutes, with mass spectrometry instrumentation typically available in hospital pathology laboratories. Prognostic tests, whilst challenging due to the varied phenotypes that may present themselves, ^9^ could be used to manage demand for hospitalisation and treatment, especially should vaccine escape lead to future waves of COVID-19 infection.

Metabolic biomarkers in serum have been identified that carry prognostic information, ^10,11^ but sampling blood is invasive. Our experience in collecting and analysing patient samples is that saliva samples are significantly easier to collect and handle than blood. Blood collection requires trained phlebotomists, causes discomfort to patients and must be spun soon after collection to preserve the metabolome. In contrast, a saliva sample can be donated quickly and painlessly by a patient. Saliva is itself a carrier of the coronavirus, ^12^ and has been proposed as a gold standard for SARS-CoV-2 detection. ^13,14^ It additionally offers information via its own characteristic metabolites. ^15^ To date, saliva as a biofluid for metabolism analysis has been used for breast, pancreatic and also oral cancers. ^16,17^ Here we explore the potential of saliva metabolomics to distinguish between severe and mild COVID-19 infection, with a view to providing a prognostic test that can be used to triage hospital patients, for example to identify patients who would benefit from immunomodulating drugs such as Tocilizumab. ^18^

This work took place as part of the wider efforts of the COVID-19 International Mass Spectrometry (MS) Coalition. ^19,20^ This consortium aims to provide molecular level information on SARS-CoV-2 in infected humans, in order to better understand, diagnose and treat cases of COVID-19 infection. Data related to this work will be stored and fully accessible on the MS Coalition open repository. The website URL is https://covid19-msc.org/

## 2. Materials and Methods

### 2.1 Participant recruitment and ethics

Ethical approval for this project (IRAS project ID 155921) was obtained via the NHS Health Research Authority (REC reference: 14/LO/1221). 88 participants were recruited at NHS Frimley NHS Foundation Trust hospitals by researchers from the University of Surrey. Participants were identified by clinical staff to ensure that they had the capacity to consent to the study, and were asked to sign an Informed Consent Form; those that did not have this capacity or who did not sign the form were not sampled. Consenting participants were categorised by the hospital as either “query COVID” (meaning there was clinical suspicion of COVID-19 infection) or “COVID positive” (meaning that a positive COVID test result had been recorded during their admission). All participants were provided with a Patient Information Sheet explaining the goals of the study.

Inclusion for participants was determined by reverse transcription polymerase chain reaction (RT-PCR) results; participants with an inconclusive RT-PCR test (clinically positive only and/or inconclusive test result, n=6) or where the time lag between initial RT-PCR test and sampling exceeded fourteen days were excluded (n=7). These additional exclusion criteria reduced the participant population from 88 to 75.

### 2.2 Sample collection, extraction and instrumental analysis

Patients were sampled immediately upon recruitment to the study in two waves, one between May and August 2020 and the second between October and November 2020. The range in time between symptom onset and saliva sampling ranged from 1 day to > 1 month, an inevitable consequence of collecting samples in a pandemic situation. Each participant provided a sample of saliva by spitting directly into a falcon tube which was placed on ice immediately after collection. Samples were transferred on ice from the hospital to the University of Surrey by courier within 4 hours of collection, to minimise changes to salivary metabolites. ^21^ Once received at University of Surrey, the samples were stored at minus 80 °C until analysis.

Alongside saliva collection, metadata for all participants was also collected covering *inter alia* sex, age, comorbidities (based on whether the participant was receiving treatment), the results and dates of COVID PCR tests, bilateral chest X-Ray changes, smoking status, drug regimen, and whether and when the participant presented with clinical symptoms of COVID-19. Values for lymphocytes, CRP and eosinophils were also taken; values obtained within five days of the saliva sampling were recorded. Each participant was attributed a “severity score” in relation to their fitness observations at the time of hospital admission using the metadata collected. This score used the “mortality scoring” approach of SR Knight et *al*. ^8^ adapted to disregard age, sex at birth and comorbidities, and ranged from 0 to 6; patients scoring 0 to 3 were attributed low severity and patients scoring 4 to 6 were attributed high severity.

Sample preparation and processing followed the guidelines set out by the COVID-19 Mass Spectrometry Coalition. ^22^ Saliva samples were separated into aliquots: 50 µL of saliva was added to 200 µL of ice-cold isopropanol to precipitate protein, and this also had the advantage of deactivating the virus to allow transfer into a lower biological safety level laboratory. The samples were agitated for one hour, sonicated three times for 30 seconds, with resting on ice for 30 seconds between each sonication. Each sample was then left to stand on ice for 30 minutes then centrifuged for 10 minutes at 10 000 g. The supernatant was removed and the precipitated protein pellet reserved for future analysis. The supernatant then underwent centrifugal filtration (0.22 µm cellulose acetate) for five minutes at 10,000 g, and the filtered supernatant was then dried under nitrogen and stored at minus 80 °C.

Samples were reconstituted on the day of analysis in 100 µL water:methanol (95:5) with 0.1% formic acid by volume. 10 µL of each sample was set aside for combination into a pooled QC. The samples were analysed over a period of eleven days. Each day consisted of a run incorporating blank injections (n=2), field blank injections (n=3), pooled QC injections (n=6, 3 at the start and finish), as well as QCs to measure instrumental and extraction variation (n=7 and 3 respectively), and 10 participant samples, randomised for positive/negative, with 3 repeat analyses for each.

### 2.3 Materials and chemicals

The materials and solvents utilised in this study were as follows: 2 mL microcentrifuge tubes (Eppendorf, UK), 0.22 µm cellulose acetate sterile Spin-X centrifuge tube filters (Corning incorporated, USA), 200 µL micropipette tips (Starlab, UK) and Qsert™ clear glass insert LC vials (Supelco, UK). LC-MS grade 2-propanol was used as an inactivation solvent. Optima™ LC-MS grade methanol and water were used as reconstitution solvents and mobile phases. LC-MS grade formic acid was added to the mobile phase solvents at 0.1% (v/v). Solvents were purchased from Fisher Scientific, UK.

### 2.4 Instrumentation and operating conditions

Analysis of samples was carried out using a UltiMate 3000 UHPLC equipped with a binary solvent manager, column compartment and autosampler, coupled to a Q Exactive™ Plus Hybrid Quadrupole-Orbitrap™ mass spectrometer (Thermo Fisher Scientific, UK) at the University of Surrey’s Ion Beam Centre. Chromatographic separation was performed on a Waters ACQUITY UPLC BEH C18 column (1.7 µm, 2.1 mm x 100 mm) operated at 55 °C with a flow rate of 0.3 ml min^-1^.

Mobile phase A was water: methanol (v/v 95:5) with 0.1% formic acid, whilst mobile phase B was methanol:water (v/v, 95:5) with 0.1% formic acid (v/v). An injection volume of 5 µL was used. The initial solvent mixture was 2% B for one minute, increasing to 98% B over 16 minutes and held at this level for four minutes. The gradient was finally reduced back to 2% B and held for two minutes to allow for column equilibration. Analysis on the Q-Exactive Plus mass spectrometer was performed with a scan range of *m/z* 100 to 1 000, and 5 ppm mass accuracy. MS/MS validation of features was carried out on Pooled QC samples using data dependent acquisition mode and normalised collision energies of 30 and 35 (arbitrary units). Operating conditions are summarised in Table S1 (Supplementary Material).

### 2.5 Data processing

LC-MS outputs (.raw files) were pre-processed for alignment and peak identification using Compound Discoverer version 3.1 and Freestyle 1.6 (Thermo Fisher Scientific, UK). Peak picking was set to a mass tolerance ±5 ppm, and alignment to a retention time window of 120 seconds. Missing values were imputed using a K-nearest neighbour approach. ^23^ Features identified by mass spectrometry were initially annotated using accurate mass match with reference to external databases (KEGG, Human Metabolome Database, DrugBank, LipidMaps and BioCyc), and then validation was performed using data dependent MS/MS analysis. This process yielded an initial peak:area matrix with 10,700 discrete features. Two criteria were used for inclusion in the final analysis: only those features with identities validated by MS/MS were used, reducing the number of features to 1,874, and 1,514 features that were present in less than 30% of participant samples were excluded. This left 360 features that were used in the analysis. Normalisation was performed using EigenMS in NOREVA for each dataset analysed, ^24,25^ i.e. independently for the diagnostic population (COVID-19 positive versus negative) and prognostic population (COVID-19 positive: high severity versus low severity).

### 2.6 Statistical Analysis

PCA analyses were conducted in SIMCA (Sartorius Stedim Biotech, France). PLS-DA and additional machine learning was conducted in R Studio Version 1.3.959 and MetaboAnalyst. ^26,27^ Leave-one-out cross-validation was used for model validation test accuracy, sensitivity and specificity; variable importance in projection (VIP) scores were used to assess feature significance alongside p-values and effect sizes (fold count). Batch effects were assessed by PCA analysis of both collection batches (waves one and two) and also instrument and extraction batching by day (Figures S1 and S2, Supplementary Material), showing no clustering by batches. KEGG pathway analysis was performed using MetaboAnalyst.

In prognostic analysis, given the lack of a “gold standard” reference test for whether COVID-19 is likely to be high severity or low severity (as this depends on clinical judgement), positive percent agreement (PPA) between the generated model and a high severity clinical diagnosis was used in preference to sensitivity, which measures the detection of positive instances of a disease relative to a ground truth value. Similarly, negative percent agreement (NPA) between the model and a high severity clinical diagnosis was used in preference to specificity, which measures the absence of a disease relative to a ground truth value. In diagnostic analysis, given that RT-PCR tests were available to establish a ground truth, sensitivity and specificity values were calculated alongside diagnostic accuracy.

## 3. RESULTS

### 3.1 Population metadata overview

The study population analysed in this work included 75 participants, comprising 47 participants presenting with a positive COVID-19 RT-PCR test and 28 participants presenting without. Of the positive participants, 10 were classed as presenting with high severity COVID-19, 34 were classed as presenting with low severity COVID-19, and 3 lacked sufficient clinical information for severity scoring. A summary of the metadata is shown in Table 1.

**Table 1:**
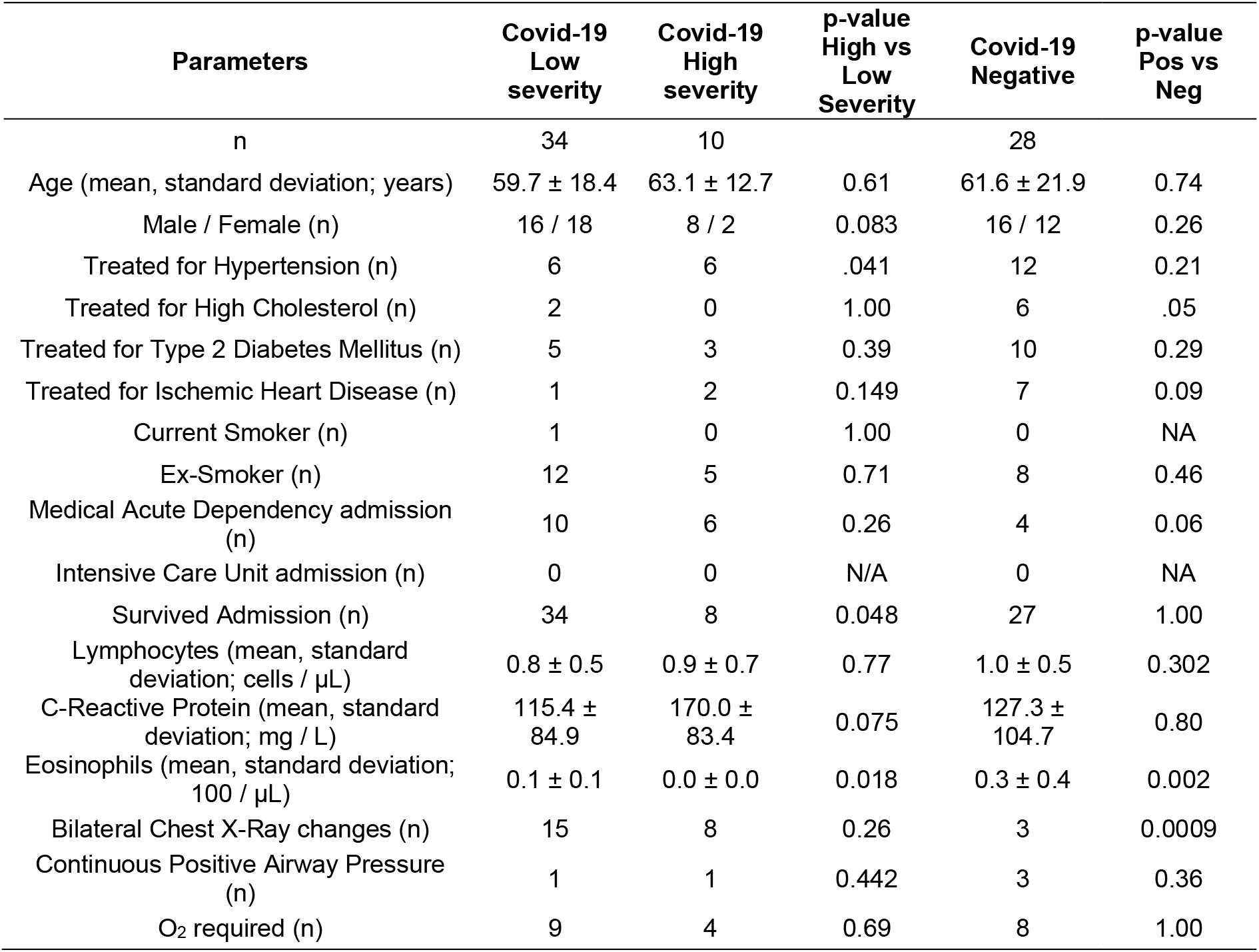
Summary of clinical characteristics by participant cohort

| Parameters | Covid-19 Low severity | Covid-19 High severity | p-value High vs Low Severity | Covid-19 Negative | p-value Pos vs Neg |
| --- | --- | --- | --- | --- | --- |
| n | 34 | 10 |  | 28 |  |
| Age (mean, standard deviation; years) | 59.7 ± 18.4 | 63.1 ± 12.7 | 0.61 | 61.6 ± 21.9 | 0.74 |
| Male / Female (n) | 16 / 18 | 8 / 2 | 0.083 | 16 / 12 | 0.26 |
| Treated for Hypertension (n) | 6 | 6 | .041 | 12 | 0.21 |
| Treated for High Cholesterol (n) | 2 | 0 | 1.00 | 6 | .05 |
| Treated for Type 2 Diabetes Mellitus (n) | 5 | 3 | 0.39 | 10 | 0.29 |
| Treated for Ischemic Heart Disease (n) | 1 | 2 | 0.149 | 7 | 0.09 |
| Current Smoker (n) | 1 | 0 | 1.00 | 0 | NA |
| Ex-Smoker (n) | 12 | 5 | 0.71 | 8 | 0.46 |
| Medical Acute Dependency admission (n) | 10 | 6 | 0.26 | 4 | 0.06 |
| Intensive Care Unit admission (n) | 0 | 0 | N/A | 0 | NA |
| Survived Admission (n) | 34 | 8 | 0.048 | 27 | 1.00 |
| Lymphocytes (mean, standard deviation; cells / μL) | 0.8 ± 0.5 | 0.9 ± 0.7 | 0.77 | 1.0 ± 0.5 | 0.302 |
| C-Reactive Protein (mean, standard deviation; mg / L) | 115.4 ± 84.9 | 170.0 ± 83.4 | 0.075 | 127.3 ± 104.7 | 0.80 |
| Eosinophils (mean, standard deviation; 100 / μL) | 0.1 ± 0.1 | 0.0 ± 0.0 | 0.018 | 0.3 ± 0.4 | 0.002 |
| Bilateral Chest X-Ray changes (n) | 15 | 8 | 0.26 | 3 | 0.0009 |
| Continuous Positive Airway Pressure (n) | 1 | 1 | 0.442 | 3 | 0.36 |
| O<sub>2</sub> required (n) | 9 | 4 | 0.69 | 8 | 1.00 |

In this study all participants were recruited in a hospital setting with at least potential suspicion of COVID-19 infection; controls were age matched and had similar profiles in terms of gender, oxygen requirements and survival rates. Significantly more COVID-19 positive patients had bilateral chest X-ray changes (p-value 0.0009) and higher levels of eosinophils (p-value 0.002), in agreement with literature observations, ^10^ but not for C-reactive protein (CRP, p-value 0.80). Type 2 diabetes mellitus (T2DM) was more prevalent in the COVID-19 negative population than the positive population, being observed in 36% of COVID-19 negatives versus 30% of high severity COVID-19 patients and 15% of low severity COVID-19 patients, and a similar observation of greater comorbidity being seen in the negative population was also true for ischemic heart disease (IHD) and hypertension (HTN). The greater preponderance of underlying comorbidities within the negative population represents a confounding factor.

Within the COVID-19 positive cohort, comorbidities were again age matched, but the high severity grouping had more males (80% male for high severity versus 47% for low severity) and had a statistically significant difference in proportion presenting with hypertension (p-value 0.04) and a statistically significant decrease in eosinophil levels (p-value 0.02). CRP was increased by a 1.5x fold count in high severity participants versus low (p-value 0.08). There was no statistically significant increase in CRP for low severity versus COVID-19 negative participants, but this may reflect changes to the inflammatory response caused by interventions reducing CRP levels in cases of mild COVID-19.

### 3.2 Overview of features identified by Liquid Chromatography Mass Spectrometry (LC-MS)

360 features with MS/MS validation were identified as being present in 30% or more of participant samples. Of these 360 features, 37 were identified as related to medical interventions or food and were excluded, leaving 323 for statistical analysis. Of the 323, 38 were annotated by *m/z* value, 171 were annotated by formula (elemental composition), and 114 were annotated as metabolites.

### 3.3 Analysis of cohorts by multivariate techniques

Initially separation of COVID-19 positive versus negative participants was tested, as well as separation of COVID-19 high severity and low severity. As shown in Figure 1A, separation for diagnostic purposes showed no clear separation by visual inspection and delivered R2Y of 0.78 and Q2Y of 0.18. Leave-one-out cross-validation (LOOCV) provided sensitivity of 0.74 (95% confidence interval of 0.60 - 0.86) and specificity of 0.75 (0.55 - 0.89), which was considered insufficient to justify further investigation. The most significantly dysregulated identified metabolites (measured by p-value) between positive and negative COVID-19 status are listed in table S2 (Supplementary Material).

**Figure 1:**
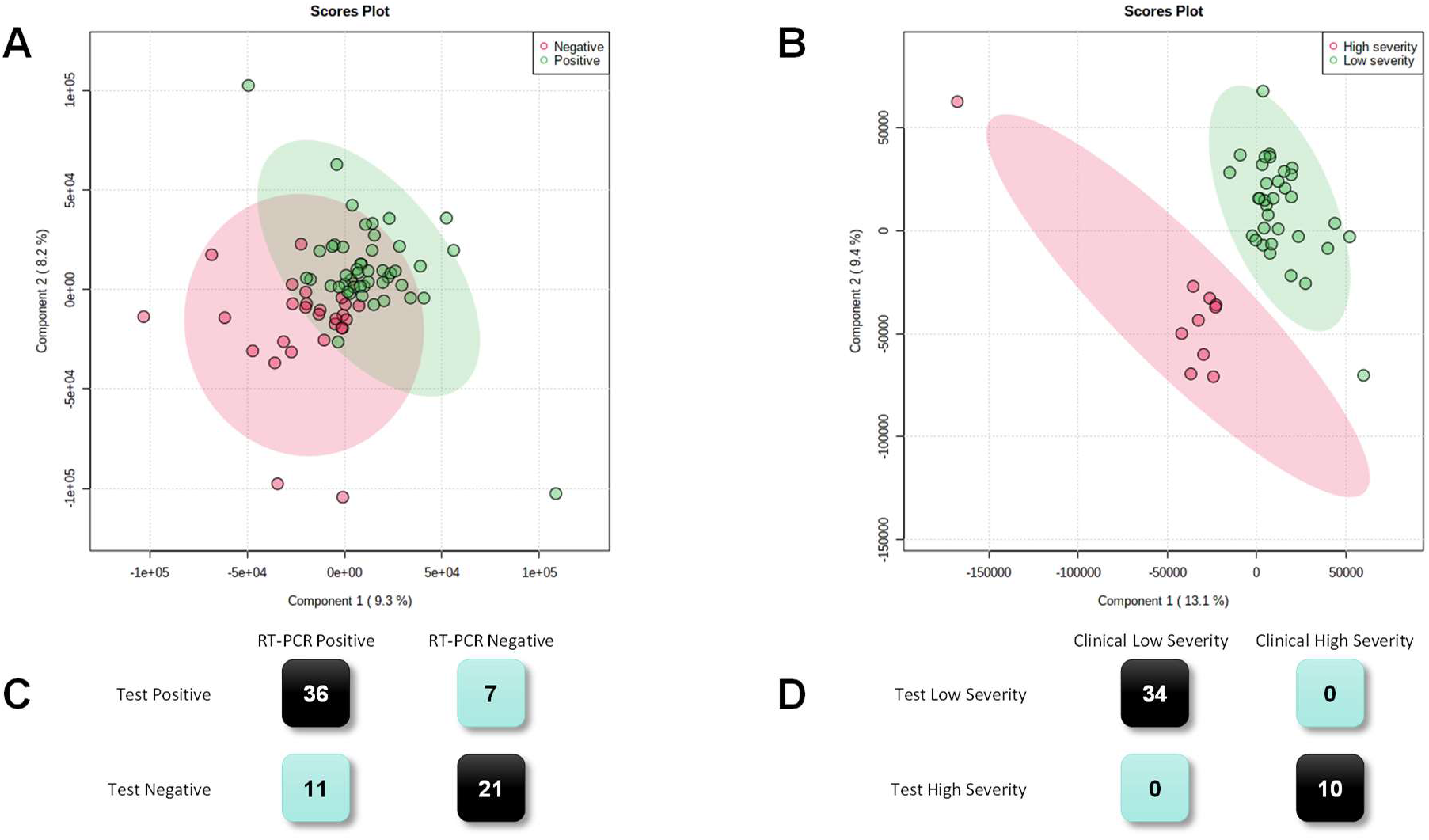
Saliva metabolomics analysis for COVID-19 diagnosis and prognosis via LC-MS, showing: **A** PLS-DA plot for 75 participants, COVID-19 positive / negative **B** PLS-DA plot for 44 participants, high severity / low severity **C** LOOCV confusion matrix, COVID-19 positive / negative **D** LOOCV confusion matrix, high severity / low severity

Figure 1B shows separation for COVID-19 high severity participants versus low severity participants. The optimal separation was found using 5 components. Using leave-one-out cross validation, PPA for COVID-19 high severity was 1.00 (95% confidence interval of 0.69 - 1.00) and NPA was 1.00 (0.90 - 1.00), for overall percent agreement with the clinical diagnosis of 1.00 (0.92 - 1.00).

A volcano plot is shown in Figure 2. The most significantly dysregulated identified metabolites (measured by p-value) are shown as boxplots in Figure 3 below and a complete list of metabolites showing statistically significant differences between high and low COVID severity populations is shown in table S3 (Supplementary Material). Amino acids are highlighted as this class of metabolites was the most dysregulated between high and low severity of the identified features.

**Figure 2:**
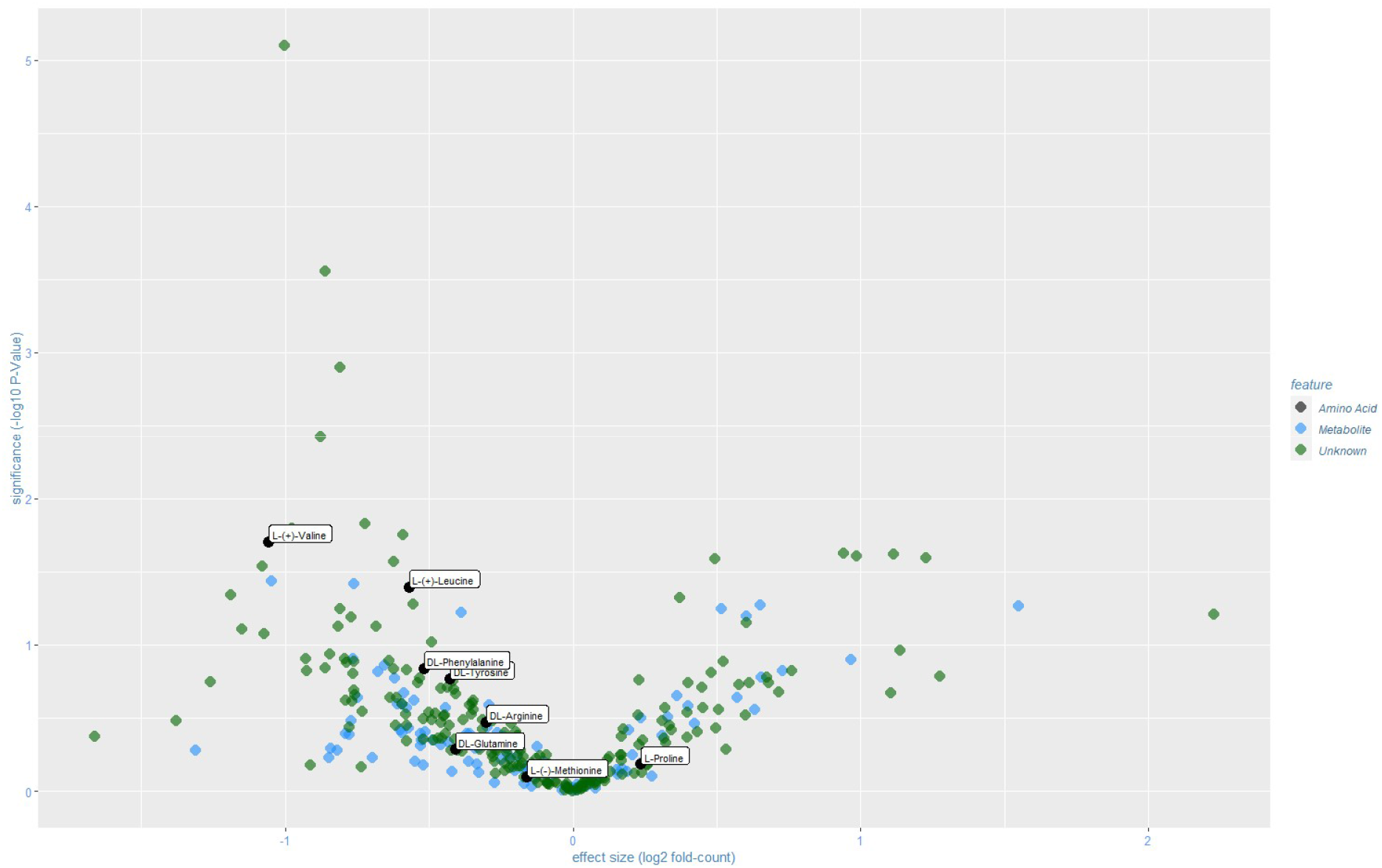
Volcano plot of statistical significance versus effect size for MS/MS validated features separating participants presenting with high severity versus low severity COVID-19

**Figure 3:**
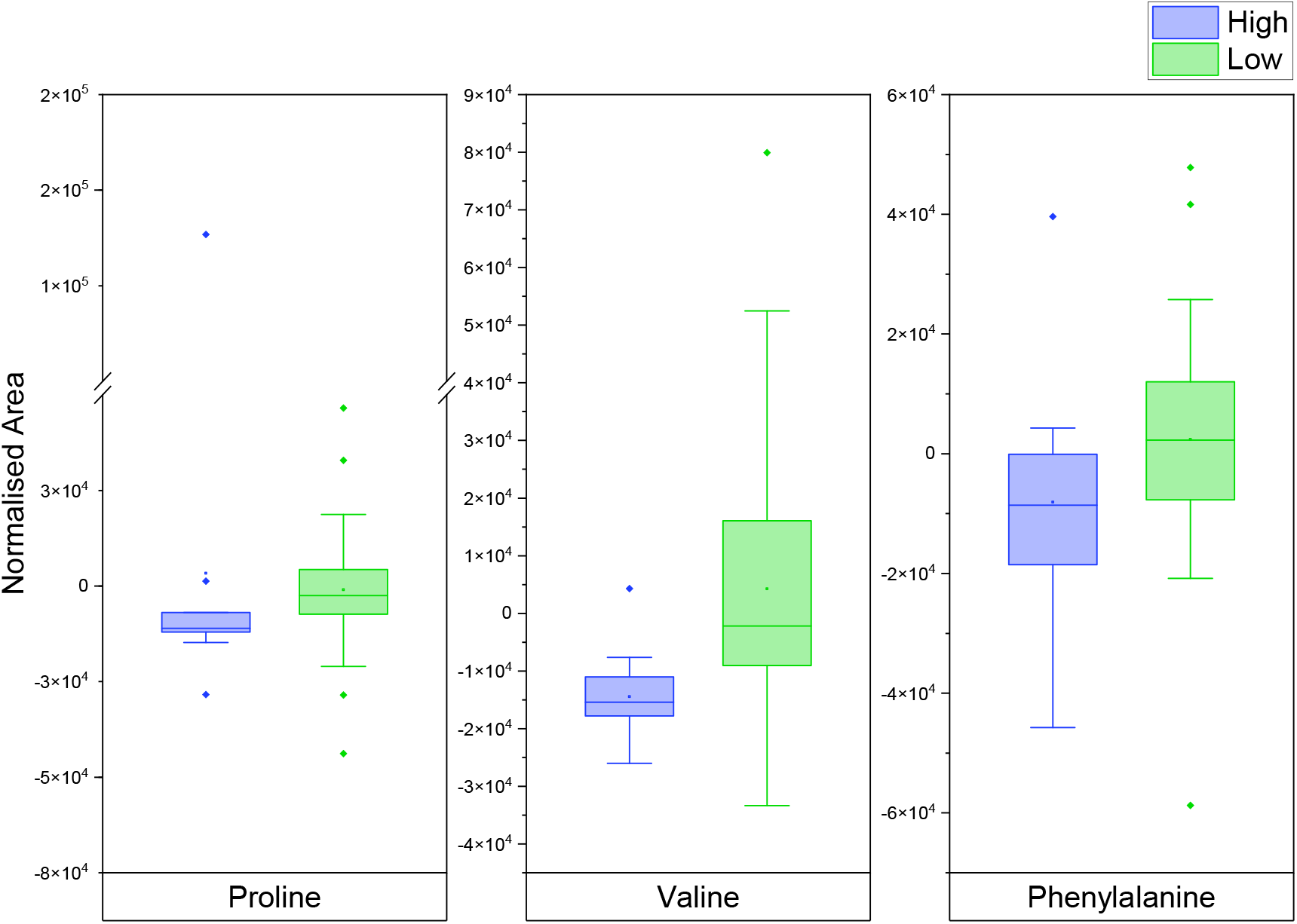
Boxplots of features down regulated with Covid-19 severity [corresponding p-values: 0.066 ; 0.004 ; 0.051]

MS/MS spectra for the significant features presented in Figure 3 are additionally shown in Figures S3 to S5 (Supplementary Material).

The normalised prognostic dataset was also processed for pathway analysis to explore changes in metabolic pathways relating to COVID-19 (Figure S6). No pathways met the criteria for both meaningful impact and statistical significance, possibly due to the number features identified in each pathway being notably smaller than typically achieved in serum or plasma, consistent with saliva being a filtrate and in general featuring lower metabolic concentrations. ^28^

### 3.4 Validation set

Whilst no fully independent prognostic validation set was available, it was decided to project the PLS-DA model obtained for high severity versus low severity participants on to COVID-19 negative participants. Given that these participants should not show features associated with high severity COVID-19, this was considered to offer additional information. The confusion matrix for the results of the projection is shown in Table 2 below.

**Table 2:** Confusion matrix for PLS-DA model projected on to COVID-19 negative participants

|  | COVID-19 negative participants |
| --- | --- |
| PLS-DA model result: High Severity | 1 |
| PLS-DA model result: Low Severity | 27 |

## 4. Discussion

Whilst age and recruitment venue were well matched (all participants were recruited in a hospital setting including controls), a number of variables within the metadata illustrate the natural difficulties in experimental design experienced during a pandemic. Age ranges of participants were large, a wide range of comorbidities were present, and the time between symptom onset and saliva sampling ranged from 1 day to > 1 month. Participant recruitment of the most severely affected was limited by ethics approval only covering patients who could give informed consent, thereby precluding the participation of patients with the highest COVID severity. Furthermore, given the small *n* in this pilot study, precision was necessarily low and confidence intervals wide.

In this study, saliva samples were provided under conditions that could be practically achieved in a hospital pandemic setting, albeit this meant no scope for abstinence from food and / or drink before saliva sampling, and no prior rinsing of the mouth, leading to potential confounding factors. Separation of COVID-19 positive COVID-19 versus negative participants was limited, possibly due to COVID-19 negative participants also being hospitalised and in poor health, perhaps having similar inflammatory responses to some COVID-19 positive participants. In spite of this, superior differentiation by multivariate analysis was achieved in relation to COVID-19 severity. PLS-DA showed separation of High Severity COVID-19 positive participants from Low Severity COVID-19 positive participants, with PPA and NPA of 100% by LOOCV. Furthermore, whilst not a true independent validation set, projecting the PLS-DA model on to COVID-19 negative participants, i.e. the controls, showed that the model classified them with 97% consistency as “low risk”, i.e. that the features associated with high severity were present neither in low severity nor in COVID-19 negative participants.

A number of identified metabolites showed statistically significant differences between the high and low severity participants. Both valine (p-value 0.02, fold-count 0.48) and leucine (p-value 0.04, fold-count 0.67) showed statistically significant changes between high and low severity. As shown in Figure 3, amino acids constituted the class of metabolites seeing the most change between high and low severity, similar to literature observations of changes in either amino acids or ratios of amino acids, albeit specific amino acids commonly cited in the literature (for example kynurenine, arginine or ratios thereof) ^1,29,30^ did not feature in the saliva analysis presented here. It should be noted, however, that the correlation of metabolites between saliva and blood has previously been found to be weak or in some cases non-existent, ^31,32^ and the same may be true for the saliva and blood of individuals testing positive for COVID-19. Direct analysis of paired blood samples would be required to draw any definitive conclusion on differential dysregulation of metabolites between serum and saliva.

## 5. Conclusion

In this work a number of features have been identified for the first time that may differentiate the saliva of those presenting with high severity and low severity COVID-19. We believe that saliva has potential to add to understanding of the progression and severity of COVID-19. In addition, saliva may be collected less invasively than other biofluids, and mass spectrometry techniques have the advantage of being often located within hospitals, making MS-based techniques useful in a clinical setting. Consequently, we view saliva as a worthy biofluid for consideration for prognostic testing.

## Data Availability

Data related to this work will be stored and fully accessible on the MS Coalition open repository. The website URL is https://covid19-msc.org/

## Data sharing statement

Participant metadata data with identifiers, alongside peak:area matrices used in this work will be made available on the Mass Spectrometry Coalition website upon publication of this study. The analytical protocols used as well as mass spectrometry .raw files, sample and participant data will be openly available for all researchers to access. The website URL is https://covid19-msc.org/

## Author’s contributions

CF and KL collected and extracted all samples used in this work, were responsible for LC-MS method development, data processing and statistical analysis in SIMCA. MS conducted machine learning and pathway analyses and additionally drafted the manuscript. CC and HL supported mass spectrometry method development and advised on analytical methods. AS and DDW obtained ethical approval and biobanking of samples. GE and DG facilitated access to participants and collected participant metadata. DS, DT, PB, KH and AP supported experimental and statistical design and also provided editorial comment on the draft manuscript. MB oversaw all aspects of this work, including obtaining funding for the study, clinical access, experimental design, analysis and was responsible for supervision of the research team.

## Acknowledgements

The authors acknowledge Samiksha Ghimire from Groningen Medical School for translation of participant information sheets and consent forms into Nepalese, as well as Kyle Saunders of the University of Surrey for access to batch controls. The authors are additionally grateful to Thanuja Weerasinge (Jay), Manjula Meda, Chris Orchard and Joanne Zamani of Frimley Park NHS Foundation Trust for their help with ethics approvals and access to hospital patients.

## Supplementary Material

**Table S1:** Operating conditions of the mass spectrometer used in this research.

| Parameter | Operating condition |
| --- | --- |
| Spray voltage | 3.5 kV |
| Capillary temperature | 275 °C |
| S-lens RF level | 50 |
| Sheath gas flow rate | 40 |
| Aux gas flow rate | 0 |
| Scan range | 100 *m/z* to 1 000 *m/z* |
| Resolution | 70 000 |
| Polarity | Positive |
| AGC target | 10<sup>6</sup> |
| Maximum inject time | 200 |
| MS/MS Parameter | Operating condition |
| Mode | Full Scan MS / dd-MS<sup>2</sup> with inclusion lists |
| Resolution | 35 000 at *m/z* 200 |
| Loop Count | 6 |
| Intensity threshold | 5 * 10<sup>4</sup> |
| Collision energy | 30 and 35 |
| Dynamic Exclusion | 5 seconds |

**Figure S1:**
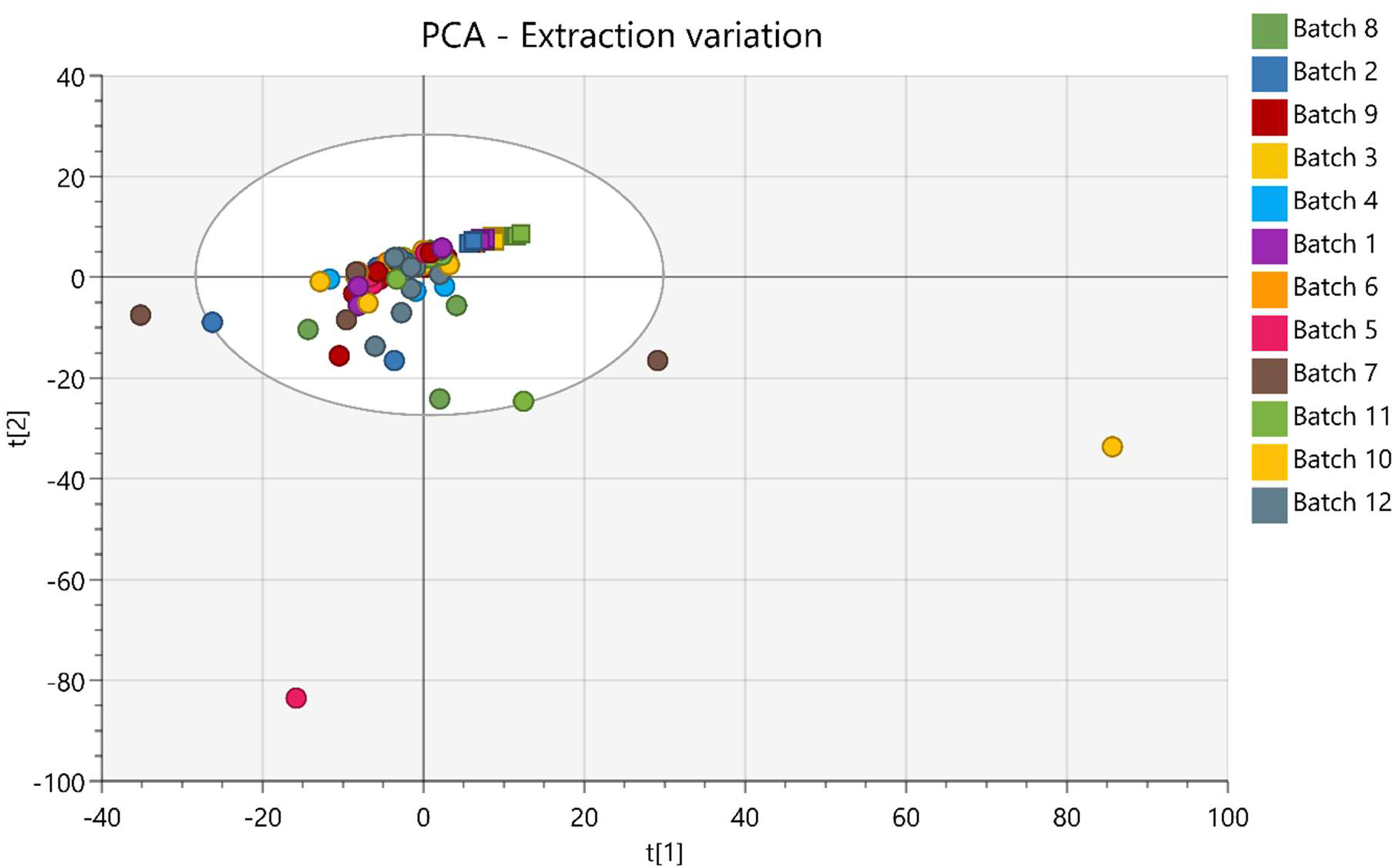
Principal Component Analysis of each patient sample (circles) and batch QC’s (squares), coloured according to extraction batch, showing no significant clustering of patient samples according to extraction batch (square: QC ; circle: patients)

**Figure S2:**
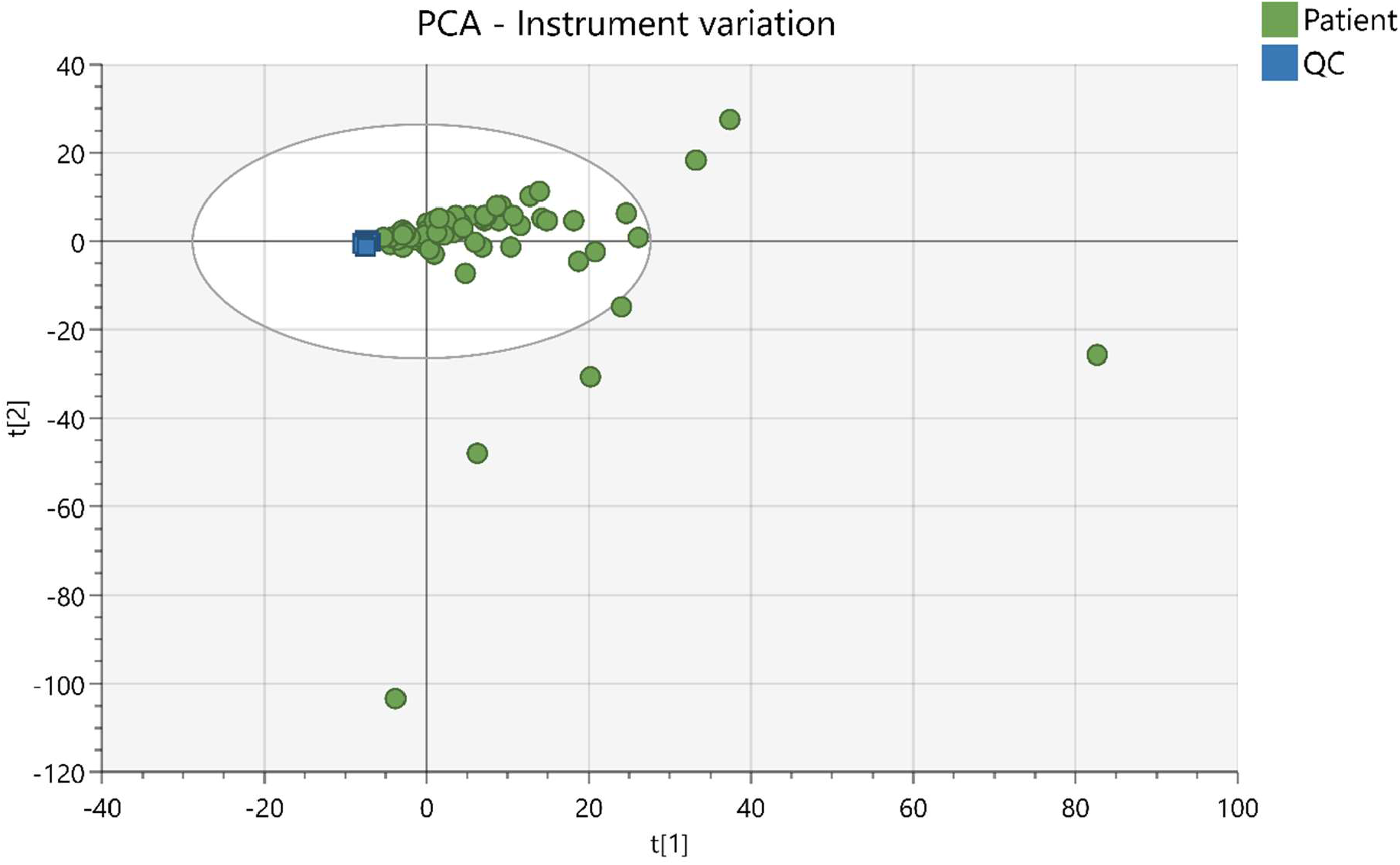
Principal Component Analysis of each patient sample and run QC, showing low levels of QC variation according to position in the run sequence

**Table S2:** Features distinctive between COVID-19 positive and negative

| Feature | Fold Count | p-value |
| --- | --- | --- |
| C25 H36 N6 O6 | 0.52 | 0.0121 |
| DL-Phenylalanine | 1.39 | 0.0164 |
| C7 H10 N6 O2 | 1.35 | 0.0179 |
| 2-linoleoyl-sn-glycero-3-phosphoethanolamine | 1.85 | 0.0180 |
| C39 H71 N2 O16 P3 | 0.81 | 0.0226 |
| DC2810000 | 1.60 | 0.0231 |
| 179.8965 | 2.09 | 0.0251 |
| 1-Hexadecanoylpyrrolidine | 1.84 | 0.0287 |
| Taurine | 1.44 | 0.0292 |
| 2707 | 1.19 | 0.0363 |
| 148.0048 | 0.70 | 0.0379 |
| C47 H84 N9 O13 P3 | 0.73 | 0.0415 |
| C22 H36 N8 O9 | 1.39 | 0.0439 |
| 3721 | 1.70 | 0.0443 |
| butyl acrylate | 1.62 | 0.0470 |
| C2 H3 N3 O2 P2 | 1.25 | 0.0501 |

**Table S3:**
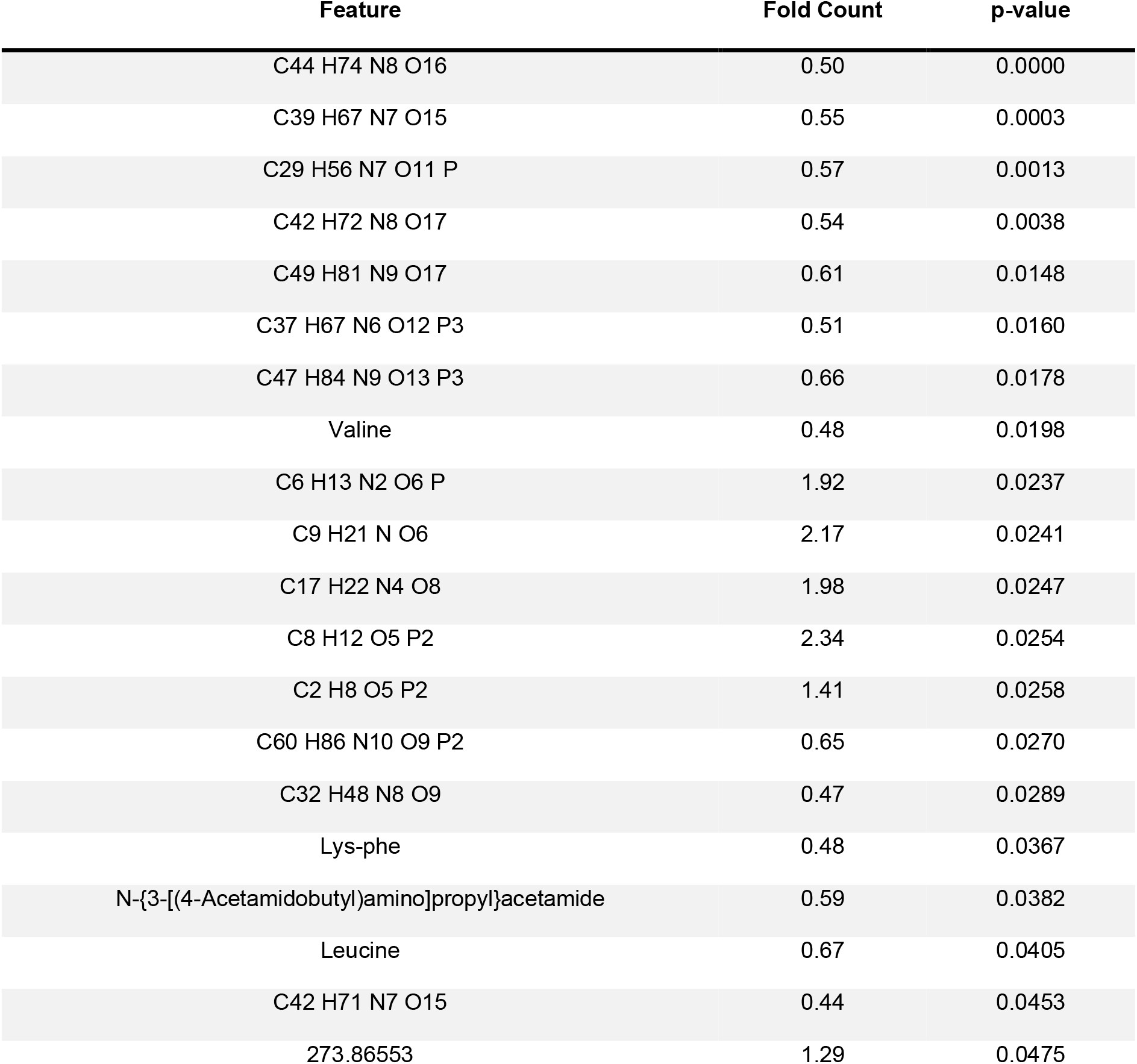
Features distinctive between COVID-19 high severity and low severity

**Figure S3:**
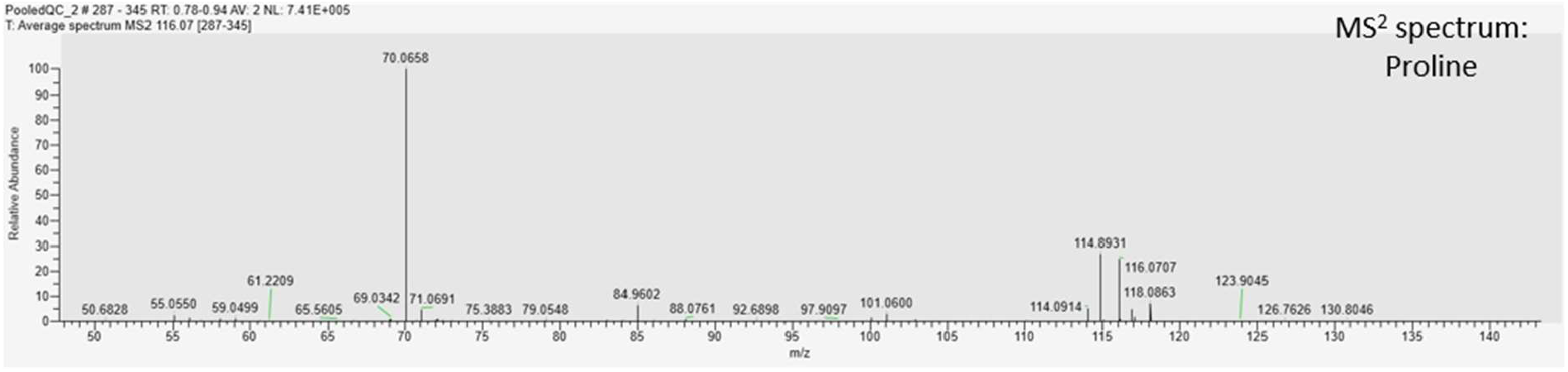
MS/MS spectra of proline

**Figure S4:**
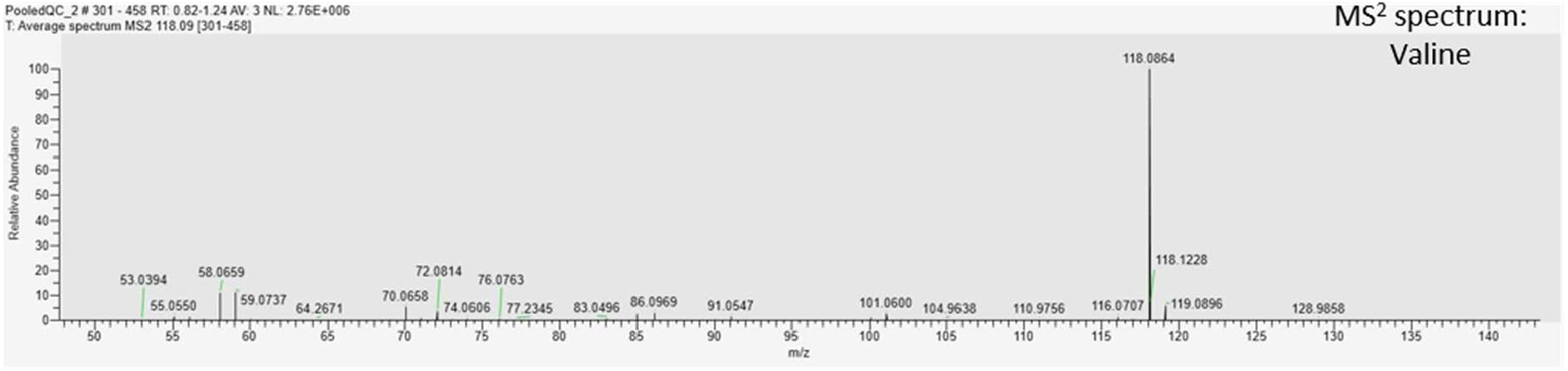
MS/MS spectra of valine

**Figure S5:**
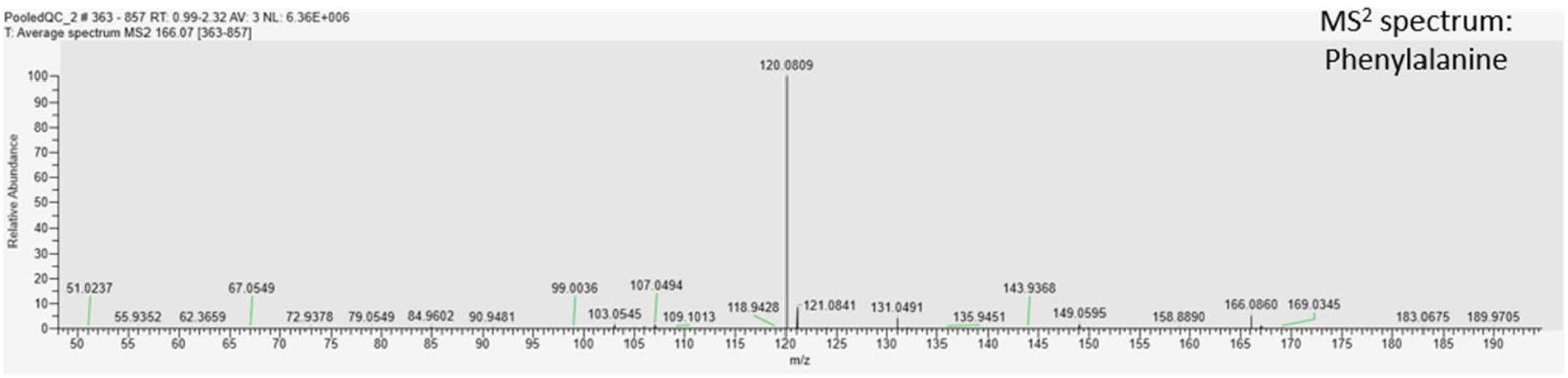
MS/MS spectra of phenylalanine

**Figure S6:**
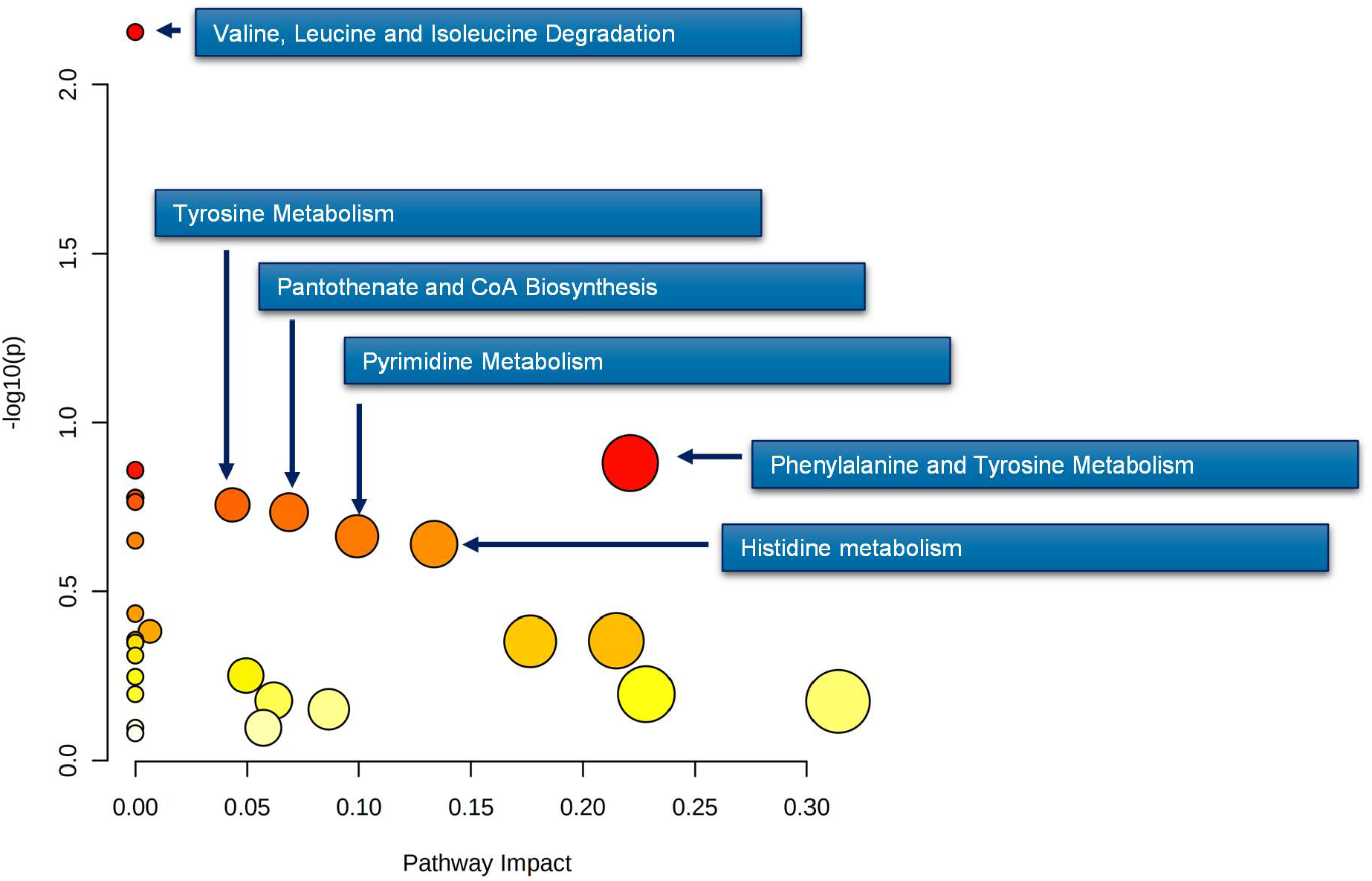
Pathways analysis of metabolites : high severity versus low severity

## Abbreviations

COVID-19: Coronavirus disease 19
CRP: C-reactive protein
HTN: Hypertension
IHD: Ischemic heart disease
KEGG: Kyoto Encyclopedia of Genes and Genomes
LC: Liquid chromatography
LC-MS: Liquid chromatography mass spectrometry
LOOCV: Leave-one-out cross validation
MS: Mass spectrometry
MS/MS or MS^2^: Tandem mass spectrometry
NPA: Negative percent agreement
PCA: Principal components analysis
PCR: Polymerase chain reaction
PLS-DA: Partial least squares-discriminant analysis
PPA: Positive percent agreement
QC: Quality control
RT-PCR: Reverse transcription polymerase chain reaction
SARS-CoV-2: Severe acute respiratory syndrome coronavirus 2
T2DM: Type 2 diabetes mellitus
VIP: Variable importance in projection

